# Interpreting Biomarker Test Results for Alzheimer’s Disease, Parkinson’s Disease and Other Neurodegenerative Diseases Without the Autopsy Gold Standard

**DOI:** 10.1101/2025.04.23.25326286

**Authors:** Nan Zhang, Charles H. Adler, Alireza Atri, Sidra Aslam, Geidy E. Serrano, Thomas G. Beach, Kewei Chen

## Abstract

There is an extensive literature on the difficulty in assessing new diagnostic tests when an accurate gold standard does not exist, is imperfect, or is not available. Relatively few reports, however, address this problem when it confronts researchers reporting tests of potential new biomarkers for Alzheimer disease (AD), Parkinson disease (PD) and other neurodegenerative diseases. This is despite the reality that the vast majority of published studies employ the neurologists’ clinical diagnoses as the gold standard, despite their well-known inherent inaccuracies relative to the true, gold standard, autopsy neuropathology diagnosis. More recently, biomarkers that have, appropriately, been evaluated against the autopsy gold standard, have then themselves been used as a surrogate gold standard to evaluate other biomarkers, despite their less-than-perfect accuracy against autopsy. The shortcomings of these approaches to neurodegenerative disease biomarker validation are rarely discussed.

It is clear from the prior literature that testing the accuracy of a new AD or PD diagnostic test against the clinical diagnosis can in fact lead to both underestimates and overestimates of its true (against autopsy) accuracy. Despite these problems, the clinical diagnoses of AD and PD are still routinely used to assess the accuracy of new biomarkers. A related issue is that when a new biomarker is evaluated against a test type previously validated against autopsy (e.g. amyloid PET tau PET), in effect serving as a surrogate gold standard, it is again clear that this new biomarker might itself be more accurate, as accurate or less accurate than the surrogate gold standard. We here present a method that makes it possible, if there are published data on the accuracy of the clinical diagnosis, or of a surrogate gold standard, relative to autopsy, to at least estimate a range of possible accuracies of a new biomarker using those imperfect gold standards. Our procedure was developed using basic theoretical modeling of conditional probabilities.

## Introduction

There is an extensive literature on the difficulty in assessing new diagnostic tests when an accurate gold standard does not exist, is imperfect, or is not available [1–7]. Relatively few reports [8], however, address this problem when it confronts researchers reporting tests of potential new biomarkers for Alzheimer’s disease (AD), Parkinson’s disease (PD) and other neurodegenerative diseases. This is despite the reality that the majority of published studies employ the neurologists’ clinical diagnoses as the gold standard, despite their well-known inherent inaccuracies [9–19] relative to the “true” gold standard autopsy neuropathology diagnosis. More recently, biomarkers that have, appropriately, been evaluated against the autopsy gold standard, have then themselves been used as a “surrogate” gold standard to evaluate other biomarkers, despite their known less-than-perfect accuracy against autopsy. The shortcomings of these approaches to neurodegenerative disease biomarker validation are rarely discussed.

Upon the introduction of a new biomarker test, it is initially unknown whether it might be more accurate, less accurate, or equivalent to, the accuracy of the clinical diagnosis. For example, an amyloid PET scan is now known to more accurately predict, as compared with the AD clinical diagnosis, the presence of clinically significant postmortem AD neuropathology [20–22]. Initial comparisons with the clinical diagnosis were puzzling, however, as up to 36% of subjects clinically diagnosed with AD dementia were found to be negative on amyloid PET scans [23], suggesting that the PET scans were poorly accurate. Subsequently, it was realized that amyloid PET is actually more accurate for predicting AD than the classical clinical diagnostic criteria.

It is clear from the literature that testing the accuracy of a new diagnostic test against the clinical diagnosis can in fact lead to both underestimates and overestimates of its true (against autopsy) accuracy. Despite this problem, the clinical diagnoses of AD and PD are still routinely used to assess the accuracy of new biomarkers. A related issue is that when a new biomarker is evaluated against a test type previously validated against autopsy (e.g. amyloid PET or tau PET), in effect serving as a surrogate gold standard, it is again clear that this new biomarker might itself be more accurate, as accurate or less accurate than the surrogate gold standard.

Statistical approaches have been developed to assist with the evaluation of diagnostic tests lacking an accurate gold standard [1–8], and we here add to this body of knowledge, especially for neurodegenerative diseases, using a simple method we derived that makes it possible, if there are published data on the accuracy of clinical diagnosis, or of a surrogate gold standard, relative to autopsy, to at least estimate a range of possible accuracies of the new test.

## Methods

The true sensitivity (Se) of a test, referred to as Test B (B for Biomarker), is a conditional probability that the test result is positive (T=1) for a patient who indeed has the disease (D=1). That is, the true sensitivity as a conditional probability is expressed as:

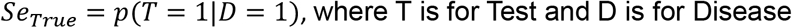

This probability *p*(*T* = 1|*D* = 1) can be decomposed to a reference standard (RS) test as true (RS=1) or as false (RS=0) in probability terms:

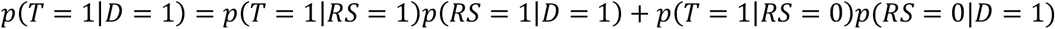

The observed sensitivity in reference to the reference standard test, is given by: *Se*_*obs*_ = *p*(*T* = 1|*RS* = 1) and the reference standard (true) sensitivity is given by: *Se*_*RS*_ = *p*(*RS* = 1|*D* = 1)

Similarly, we note, 1 − *Se*_*RS*_ = *p*(*RS* = 0|*D* = 1) and 1 − *Sp*_*obs*_ = *p*(*T* = 1|*RS* = 0) where *Sp*_*obs*_ is the observed specificity.

Putting all these together, we have expression (1):

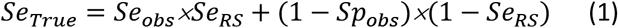

For the true specificity, *p*(*T* = 0|*D* = 0) = *Sp*_*True*_, it can be decomposed as the reference standard test as false (RS=0) or as true (RS=1) in probability terms:

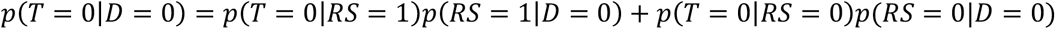

Express the reference standard specificity as *Sp*_*RS*_ = *p*(*RS* = 0|*D* = 0), and the observed specificity of Test B as *Sp*_*obs*_ = *p*(*T* = 0|*RS* = 0)

We also have 1 − *Sp*_*RS*_ = *p*(*RS* = 1|*D* = 0), and 1 − *se*_*obs*_ = *p*(*T* = 0|*RS* = 1)

Putting all these together, we get expression (2) as:

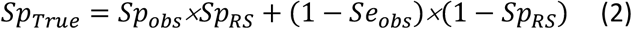

## Results

Our main results are summarized in Figures 1 and 2, each with multiple possible scenarios as subplots. For each subplot, we have two red, dashed lines. The first is the identity line (45^°^) y=x, and the second is the vertical positioned at a specified reference test sensitivity (subplots on the left side) or at a specified reference test specificity (subplots on the right side). The subplots on the left side show the relationship between the true sensitivity and observed sensitivity while the observed specificity is kept as a fixed value. The subplots on the right side show the relationship between the true specificity and observed specificity while the observed sensitivity is kept as a fixed value. In each subplot, the observed sensitivity or specificity may be located on the Y axis while its range of possible true values, located on the X axis, approximated by interpolation.

**Figure 1.**
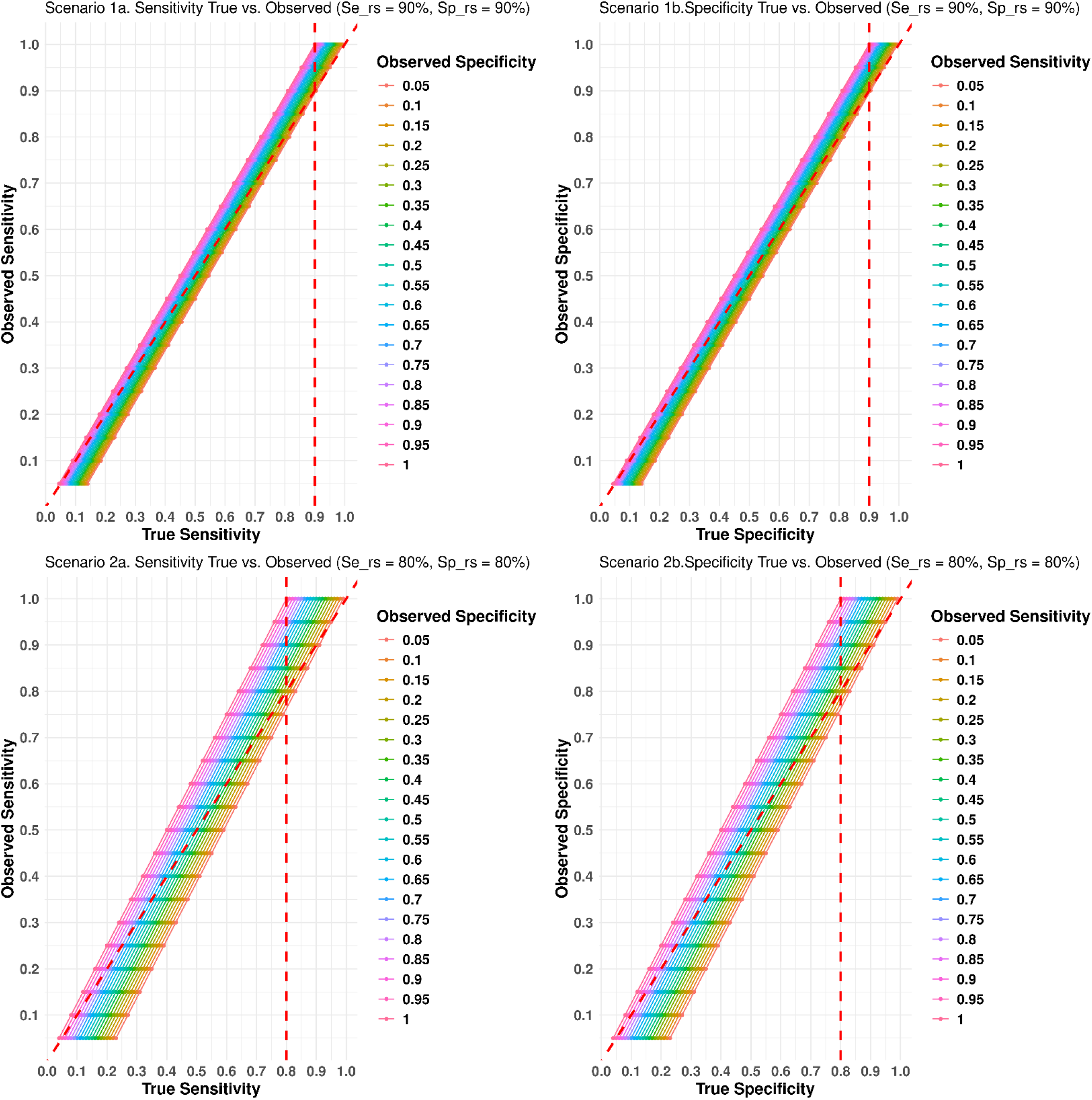
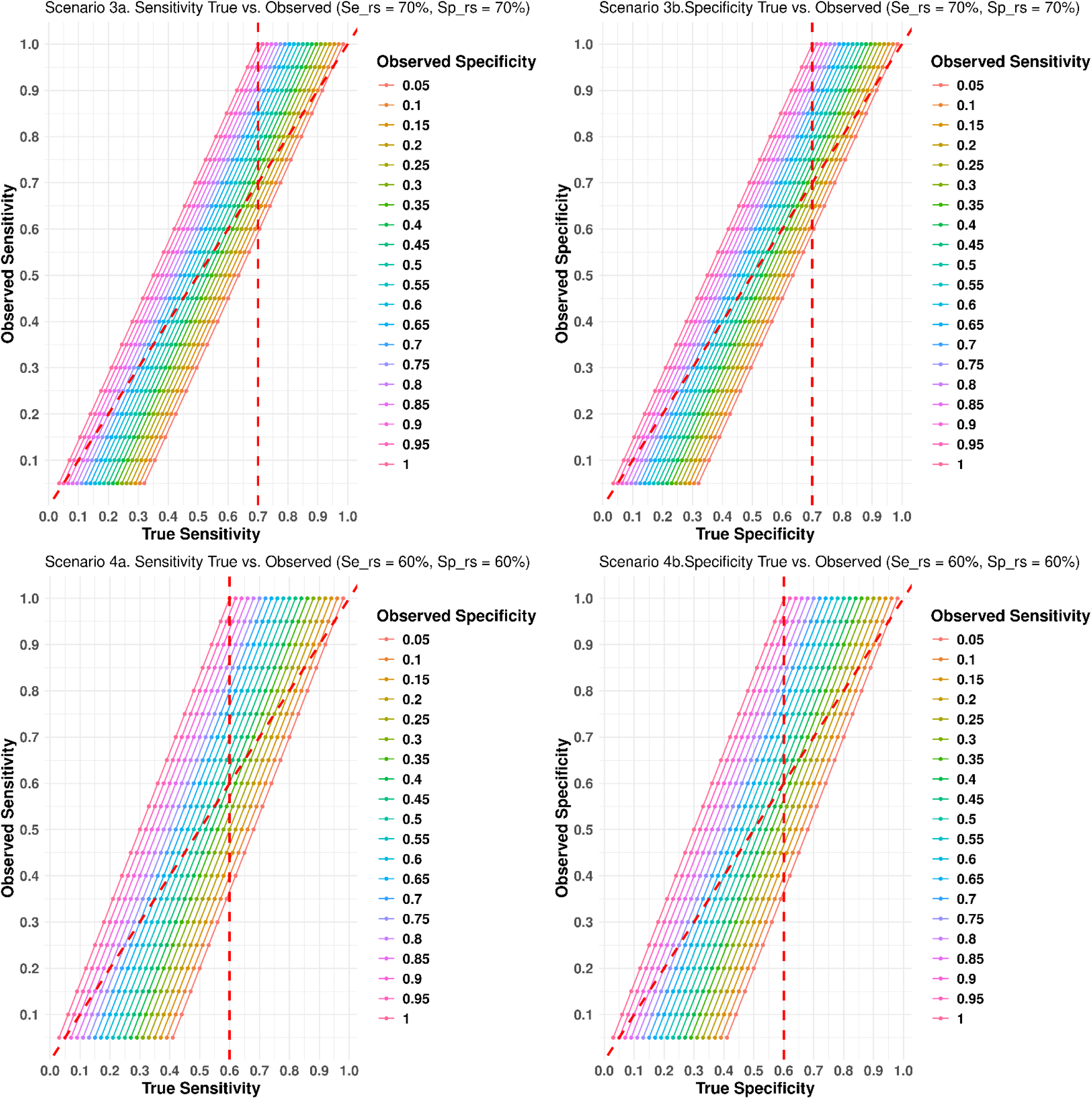
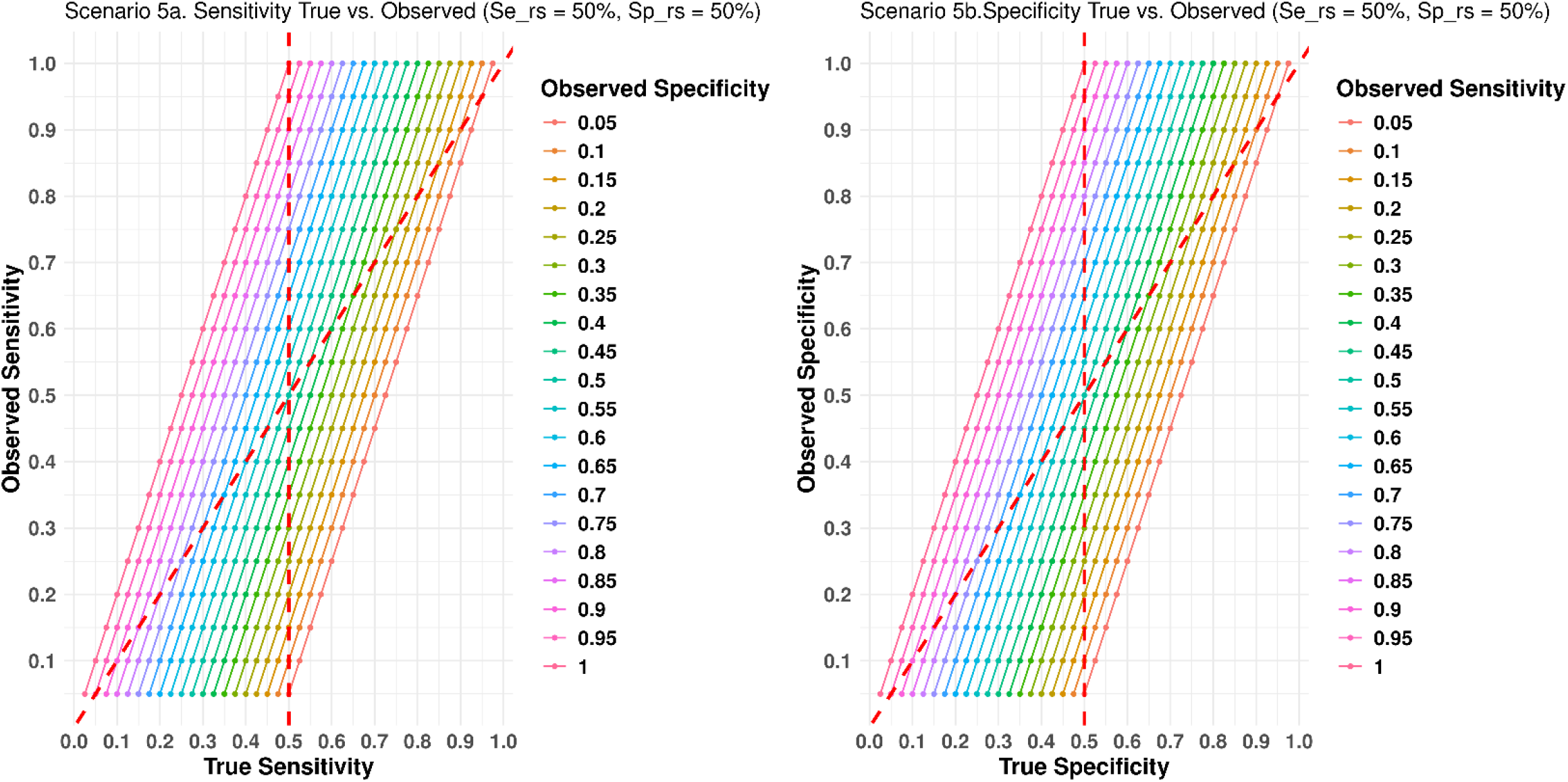
Observed versus true sensitivities and specificities with five different scenarios utilizing a hypothetical surrogate reference standard (see text for detailed explanation).

**Figure 2.**
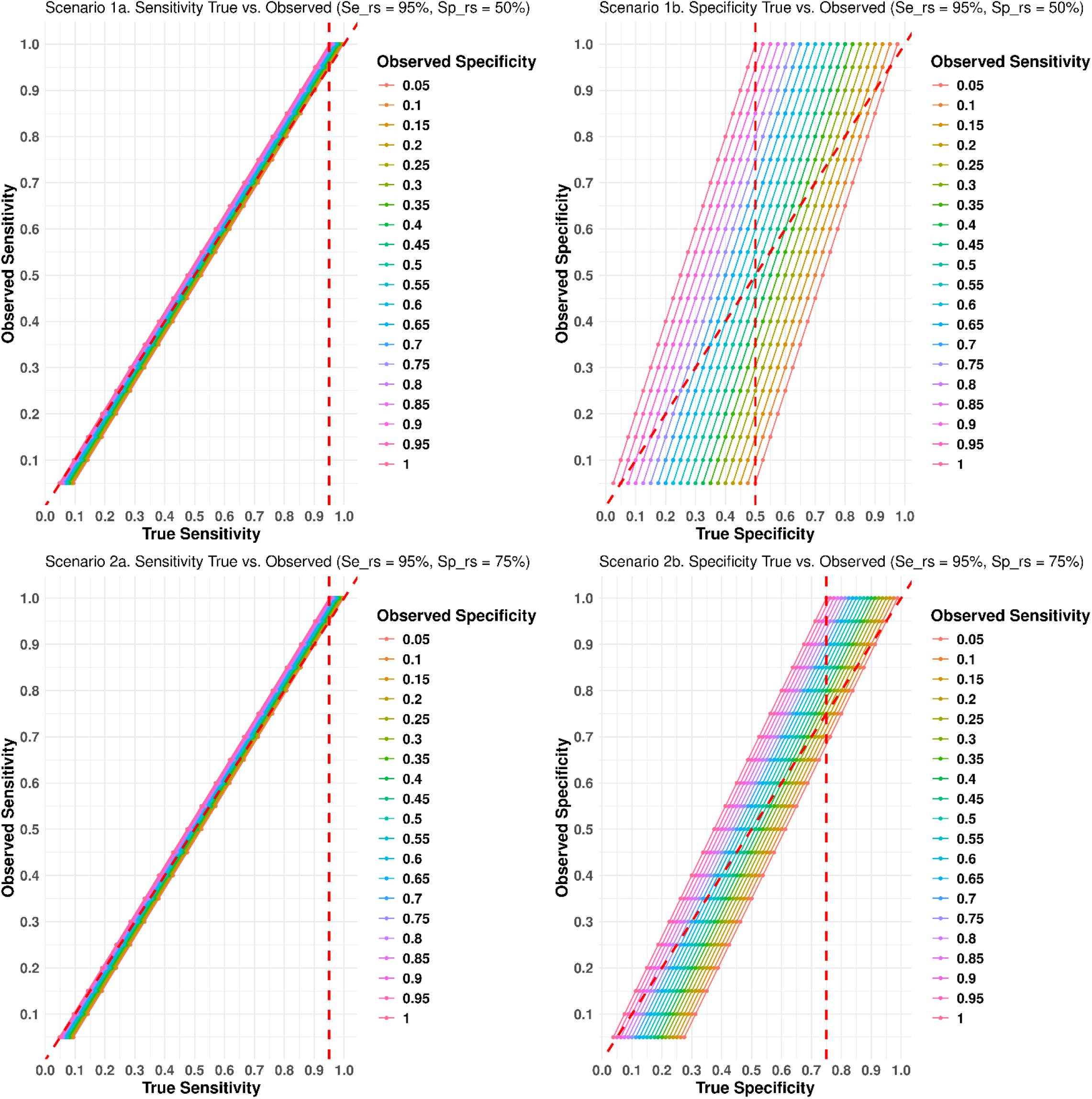
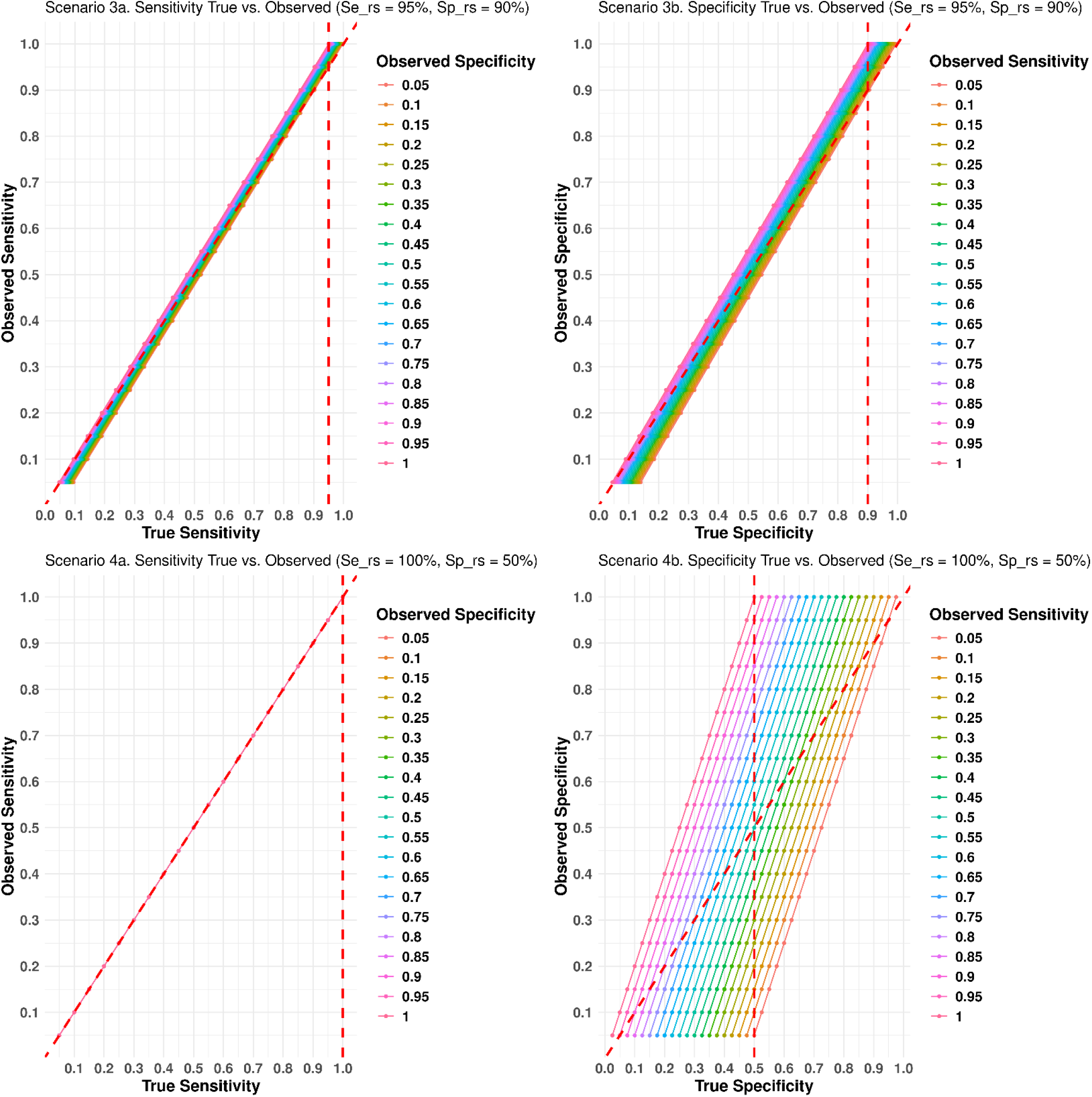
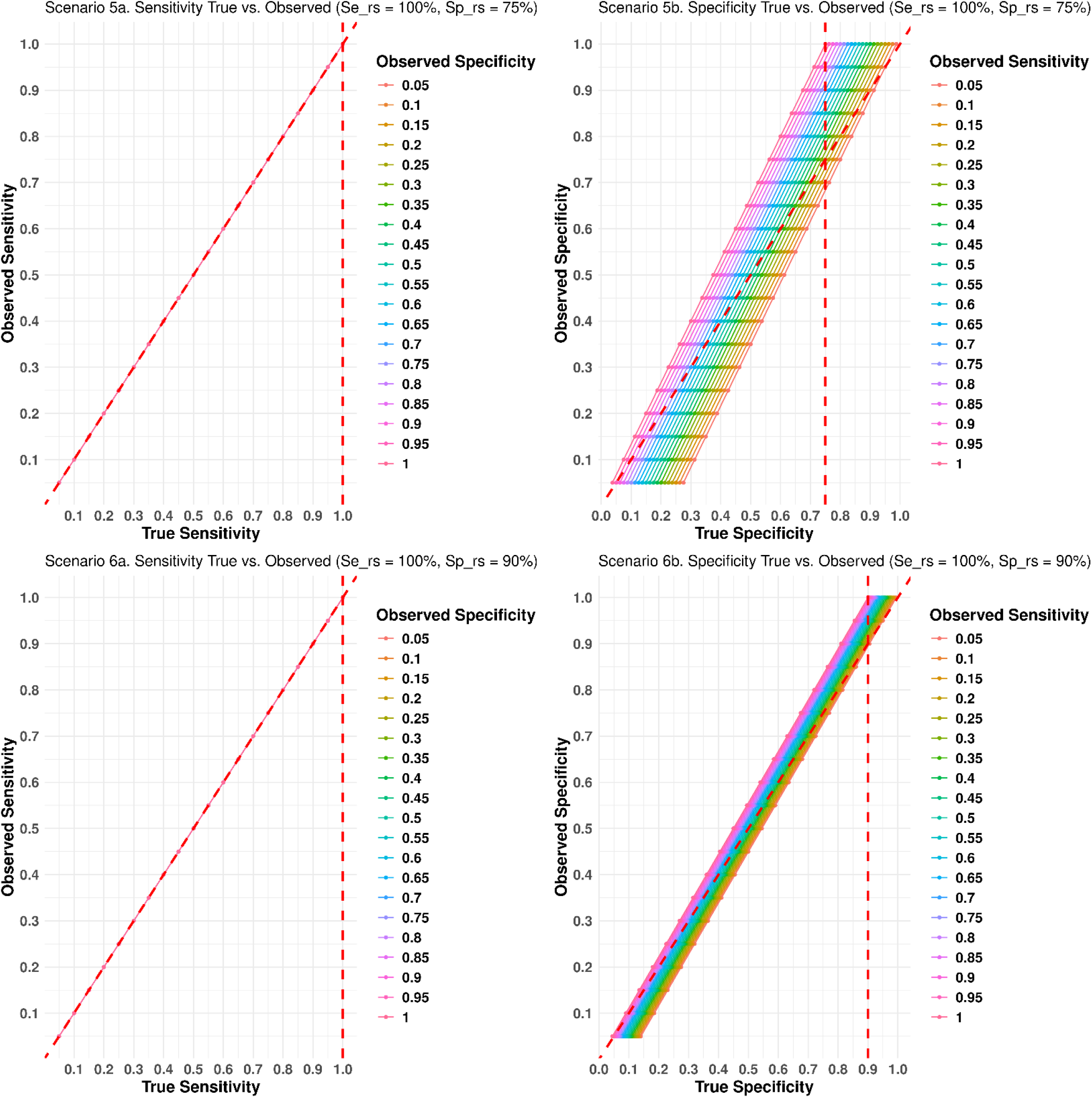
Observed versus true sensitivities and specificities for a new AD biomarker being evaluated against flortaucipir PET, with six different scenarios. See text for detailed explanation.

Based on the relationships given above, we consider the following hypothetical situations and show our results as graphically depicted below (Figure 1, Scenarios 1-5) in five possible scenarios.

In the 1^st^ scenario (Figure 1, Scenario 1a and 1b), reference sensitivity and specificity are both relatively high at 90%. In this situation, we can see the variation of the observed sensitivity and specificity is small. If the true sensitivities and specificities are greater than 90%, our observed sensitivities and specificities will overestimate the truth. The portion on the right side of the vertical line represent the overestimations as shown graphically in subplots 1a and 1b of Figure 1, all above the 45^°^ y=x identity line.

If the true sensitivities and specificities are less than 90%, then depending on different scenarios, the observed sensitivities and specificities might be either an overestimation or an underestimation, as shown in Scenarios 2-5 (Figure 1, Scenarios 2a-5a).

In Scenario 2a and 2b, the reference sensitivity and specificity are both 80%, relative to the true values. We can see that the variation, relative to the true gold standard, of the observed sensitivities and specificities is larger than in Scenario 1. If the true sensitivity (specificity) of the reference standard is greater than 80%, in most of the cases our observed sensitivity (specificity) will overestimate the true accuracy (the area on the right hand side of the 80% vertical line and above the diagonal red dashed line) but in some cases, it will underestimate the truth (the area on the right hand side of the 80% vertical line and below the diagonal red dashed line).

Scenarios 3, 4 and 5 utilize reference sensitivities and specificities of 70%, 60% and 50%, respectively. As the reference accuracy decreases, we can see that the variability of the observed sensitivity and specificity, relative to the true gold standard, increases.

In the final examples (Figure 2, Scenarios 1-6), we depict observed test results utilizing a surrogate gold standard vs the true gold standard (against autopsy), with an often-used surrogate gold standard for new AD diagnostic tests, flortaucipir (^18^F) PET. In the FDA-certifying clinical study of flortaucipir (24), it was estimated to predict an autopsy-verified “high” level of AD neuropathological change (25,26) with a sensitivity of 94.7% (95%CI, 82.7%-98.5%) to 100% (95%CI, 90.8%-100.0%) and a specificity of 50.0% (95%CI, 32.1%-67.9%) to 92.3% (95%CI, 75.9%-97.9%). New biomarker tests are often now assessed by comparing their sensitivities and specificities for the identification of subjects who have positive or negative flortaucipir PET scans. The graphs for Scenarios 1-6 in Figure 2 were generated based on this real-life scenario where flortaucipir PET reference sensitivity (for neuropathologically-classified AD) is depicted at 95% or 100% with respect to an autopsy classification of high AD neuropathological change, as well as with respect to reference specificities being equal to 50%, 75%, or 90%. We can see that the observed new biomarker test sensitivities will not be that far off from the true sensitivities. However, the observed specificities have more variability, especially when the reference specificity is as low as 50%, resulting in either overestimation or underestimation of the true specificity across a large range.

## Discussion

The majority of published studies of the accuracy of new biomarkers for AD, PD and other neurodegenerative diseases employ the neurologists’ clinical diagnoses as the gold standard, despite the well-known inaccuracies of these [9–19], relative to the true gold standard autopsy neuropathology diagnoses.

More recently, new biomarkers have been evaluated against “surrogate” gold standards that have known, still imperfect, accuracies against the autopsy gold standard. For example, amyloid PET and tau PET scans are now known to more accurately predict, as compared to the AD clinical diagnosis, the presence of clinically significant postmortem AD neuropathology [20–22], so many new AD biomarkers are now being evaluated against PET scan classifications of positive vs negative.

It is clear, from prior literature and our own analyses here, that evaluating the accuracy of a new diagnostic test against the clinical diagnosis or against a surrogate gold standard may lead to both underestimates and overestimates of the new test’s true accuracy against autopsy. Statistical approaches have been developed to assist with the evaluation of diagnostic tests lacking an accurate gold standard [1–8], and we have here added to this body of knowledge using a method we derived that makes it possible, if there are published data on the accuracy of clinical diagnosis, or of a surrogate gold standard, relative to autopsy, then it is possible to at least estimate a range of possible true accuracies of a new biomarker test, relative to its observed accuracy against the clinical diagnosis or a surrogate gold standard.

Other approaches have been made to the problem of the imperfect gold standard (27-32), in particular using latent class analysis (LCA), and its extension, Bayesian LCA. Although LCA was originally introduced to deal with the absence of a gold reference, it can also handle the “imperfectness” of existing reference standards. We introduce here a method that adjusts for an imperfect reference standard by fixing its sensitivity or specificity and estimating others relative to it. We hope that we have provided a basic understanding of the impact of an existing imperfect gold standard, targeting an audience that is interested in evaluating new biomarkers of neurodegenerative diseases.

In conclusion, we urge that researchers reporting diagnostic accuracies of new biomarkers evaluated against surrogate gold standards consider these drawbacks when interpreting their findings and consider using our formulas or graphs to calculate and report the range of possible true accuracies, relative to autopsy neuropathology.

## Data Availability

All data produced in the present study are available upon reasonable request to the authors.

